# Genomic Surveillance in Japan of AY.29—A New Sub-lineage of SARS-CoV-2 Delta Variant with C5239T and T5514C Mutations

**DOI:** 10.1101/2021.09.20.21263869

**Authors:** Takashi Abe, Masanori Arita

**Affiliations:** Faculty of Engineering, Niigata University, 8050, Ikarashi 2-no-cho, Nishi-ku, Niigata-shi, Niigata-ken 950-2181, Japan; Bioinformation and DDBJ Center, National Institute of Genetics, 1111, Yata, Mishima-shi, Shizuoka-ken 411-8540, Japan

**Keywords:** SARS-CoV-2, Delta variant (B.1.617.2), AY.29 sub-lineage

## Abstract

In the present study, we report a new sub-lineage of the SARS-CoV-2 Delta variant called AY.29, which has C5239T and T5514C mutations. We investigated the monthly trend of AY.29 in Japan within 37,737 Delta variants downloaded on October 2, 2021. Among the total Japanese Delta variants, the AY.29 sub-lineage accounted for 95.1%. In terms of monthly trends, the sequences became predominant in June, and accounted for 95.4%, 97.6% and 90.5% of the reported sequences in July, August and September, respectively. Furthermore, the number of Delta variants imported from abroad during the Tokyo 2020 Olympics and Paralympics (held in August 2021) was extremely low during the fifth wave in Japan. Therefore, the epidemic of the new Delta variant is attributable to a newly occurring mutation in Japan.

---

Severe acute respiratory syndrome coronavirus 2 (SARS-CoV-2) has spread rapidly across the globe since it was first reported in December 2019, and the momentum of its spread has not diminished (1). To address the trend of SARS-CoV-2 pandemic, genome sequencing has been performed on a global scale, and the results have been publicized at the global initiative on sharing avian influenza data (GISAID) (2). Currently, Delta variants (Phylogenetic Assignment of Named Global Outbreak (PANGO) (3) lineage: B.1.617.2) are spread worldwide and predominant in many countries including Japan after its first report in India in December 2020 (4, 5).

In the present study, genomic surveillance was performed on the genomic sequence data registered at GISAID to investigate the characteristics of the Delta variants isolated in Japan. First, to investigate the diversity, haplotype networks from genomic Delta single nucleotide variants (SNVs) were constructed using median joining network analysis (6) with PopART version 1.7 (7) against 1,348 strains downloaded on August 12, 2021 (Figure 1). The sequences were aligned using MAFFT version 7.475 (8), and sequences with gaps were extracted using trimAl version 1.4.rev22 (9). SNVs were detected using snp-sites version 2.5.1 (10), and clustered using CD-HIT version 4.8.1 (11). The resulting haplotype network was divided into five groups, described as N1–N5 in Figure 1. An examination of the hub nodes revealed that N1, N2, N3, and N5 originated from airport quarantine strains in Japan, all of which were obtained from returning travelers from India. The remaining N4 was of unknown origin. Most strains collected after June belonged to the nodes C1 to C3, generated from N5 containing Japan/IC-1040/2021. There were two mutations (C5239T and T5514C; ORF1ab: V1750A) from N5 to C1, one mutation (C28170T; ORF8: P93S) from C1 to C2, and one mutation (C5365T) from C2 to C3. Therefore, strains in C1 and C3 differed by two mutations only—i.e., C5239T and T5514C (ORF1ab: V1750A). The relationships between protein functions and mutations T5514C (ORF1ab: V1750A) and C28170T (ORF8: P93S)—which are non-synonymous substitutions—are unknown. No other mutations, except for those characterizing the Delta variant, were detected.

**Figure 1.**
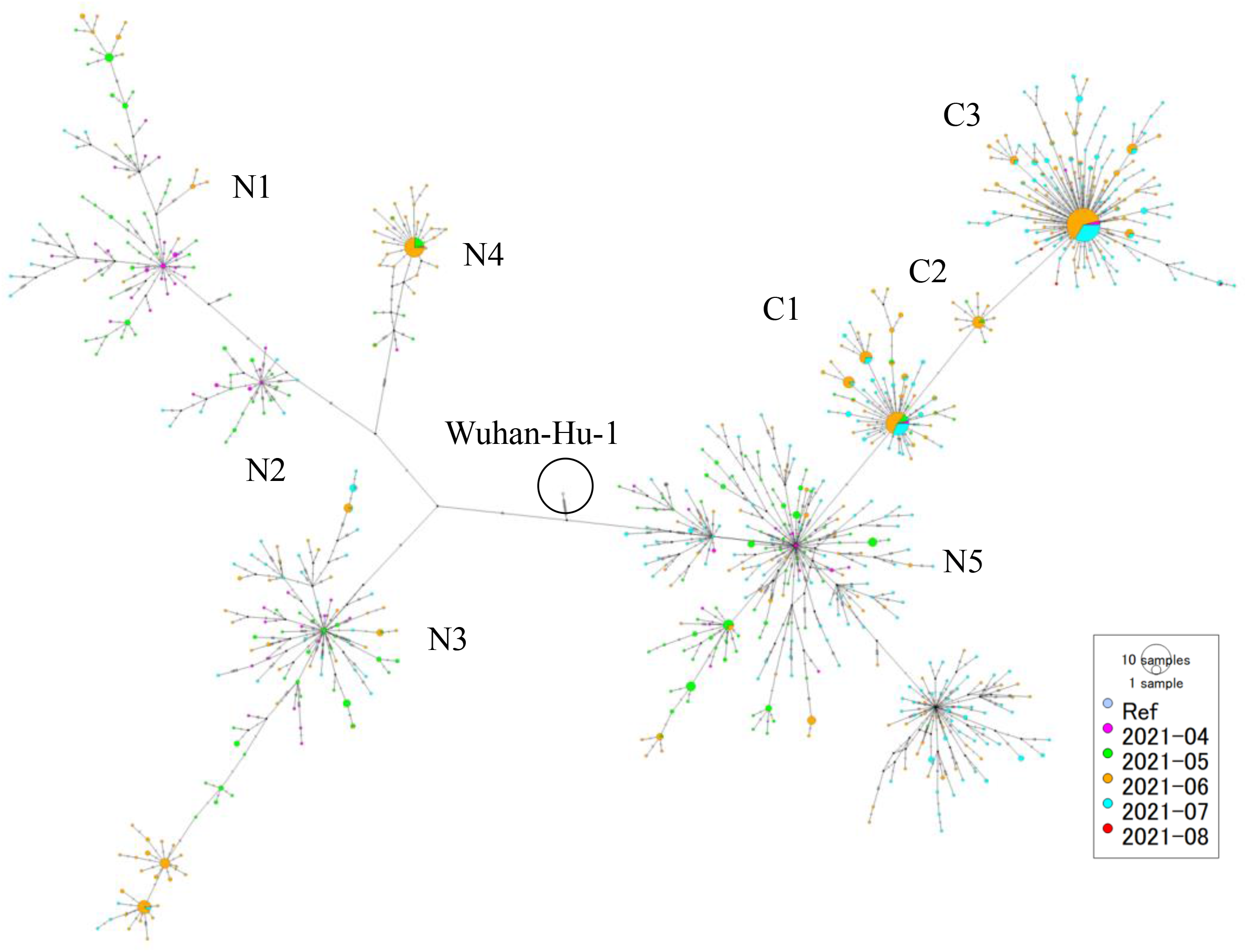
Haplotype networks. Representative groups are shown in N1 to N5. Nodes in which the strains isolated after June were mainly classified are indicated in C1 to C3. The colors indicate the months in which the strains were isolated at the bottom right of the figure. The sizes of the circles indicate the number of strains.

Next, to determine whether mutations C5239T and T5514C (ORF1ab: V1750A) are unique to Japanese strains, we searched all Delta variants downloaded on September 1, 2021. Of the 908 sequences detected with these two mutations, the countries and numbers of strains were as follows: Japan (883 strains), Hong Kong (2), Belgium (1), Italy (1), United Kingdom (13), USA (2), and New Zealand (6). Japanese strains accounted for 97.2% of the total population. The earliest strains were detected on April 2021 in Japan, and the most recent strains were detected on August 23, 2021 in the UK and Belgium. When the relationship between these strains on the phylogenetic tree was investigated, they were assigned to the new sub-lineage into the Ultrafast Sample placement on Existing tRee (UShER) global phylogenetic tree (12) (Figure 2). Therefore, mutations C5239T and T5514C (ORF1ab: V1750A) are unique to Japan.

**Figure 2.**
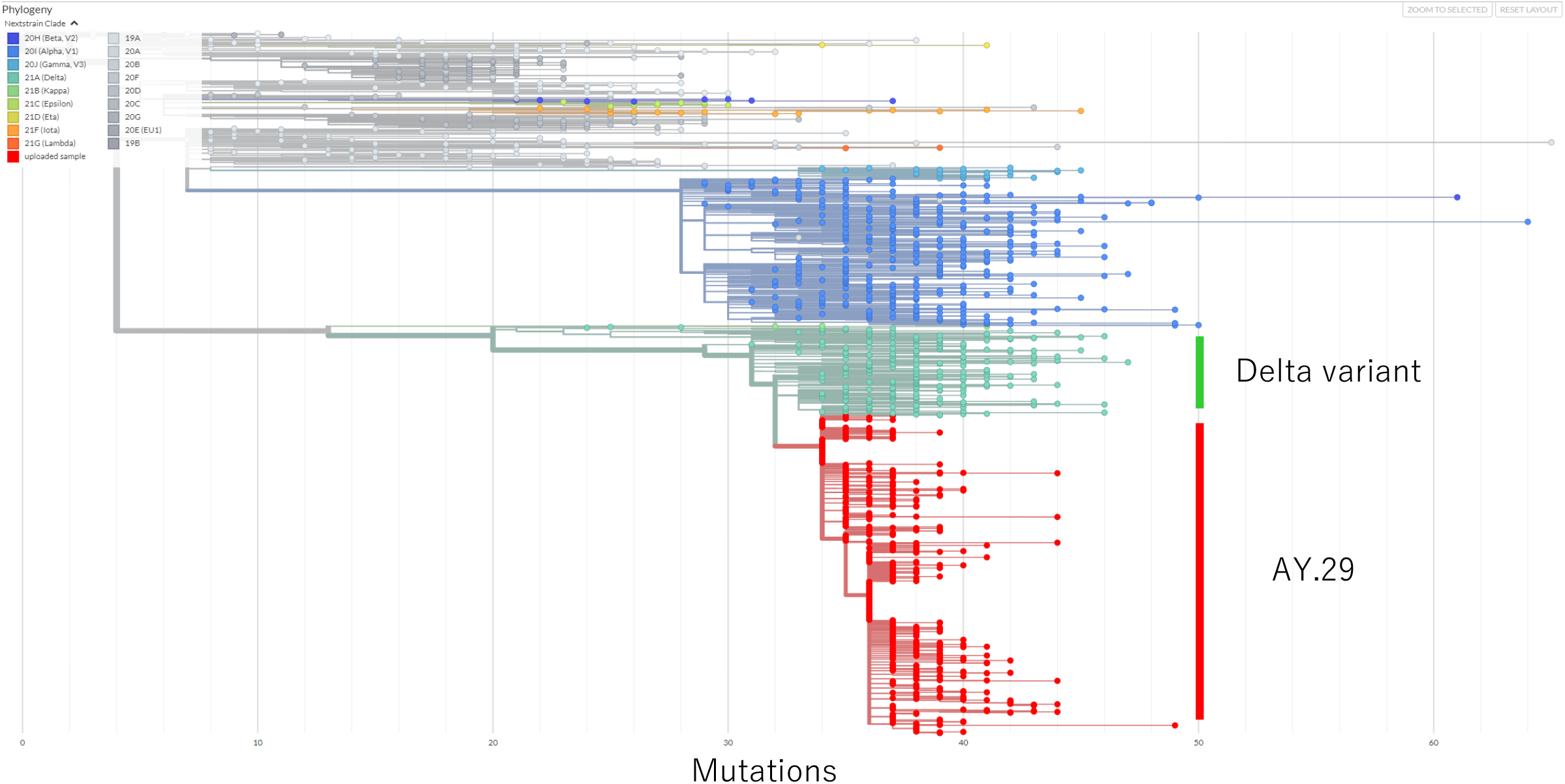
Phylogenetic tree derived from candidate sequences assigned by UShER. The colors indicate the clades. The candidate strains are shown in red, and the other strains are shown in the upper right of the figure.

Based on these results, on September 2, 2021 we proposed to the PANGO nomenclature team—which manages the genetic lineage information about SARS-CoV-2—that the strains with these mutations constitute a new sub-lineage of the Delta variant. After confirmation by PANGO, AY.29 was assigned as a new sub-lineage for Delta variants (13).

Finally, we investigated the monthly trends of AY.29 in Japan within 37,737 Japanese Delta variants downloaded on October 2, 2021 (Figure 3). Sequences derived from AY.29 accounted for 95.1% (35,904/37,737 strains). The sequences became predominant in June, and accounted for 95.4% (11,419/11,967 strains), 97.6% (23,110/23,669 strains), and 90.5% (735/812 strains) of the detected sequences in July, August and September, respectively. In addition to these two mutations, strains with C28170T (ORF8: P93S) and C5365T mutations also accounted for approximately half of the AY.29 cases.

**Figure 3.**
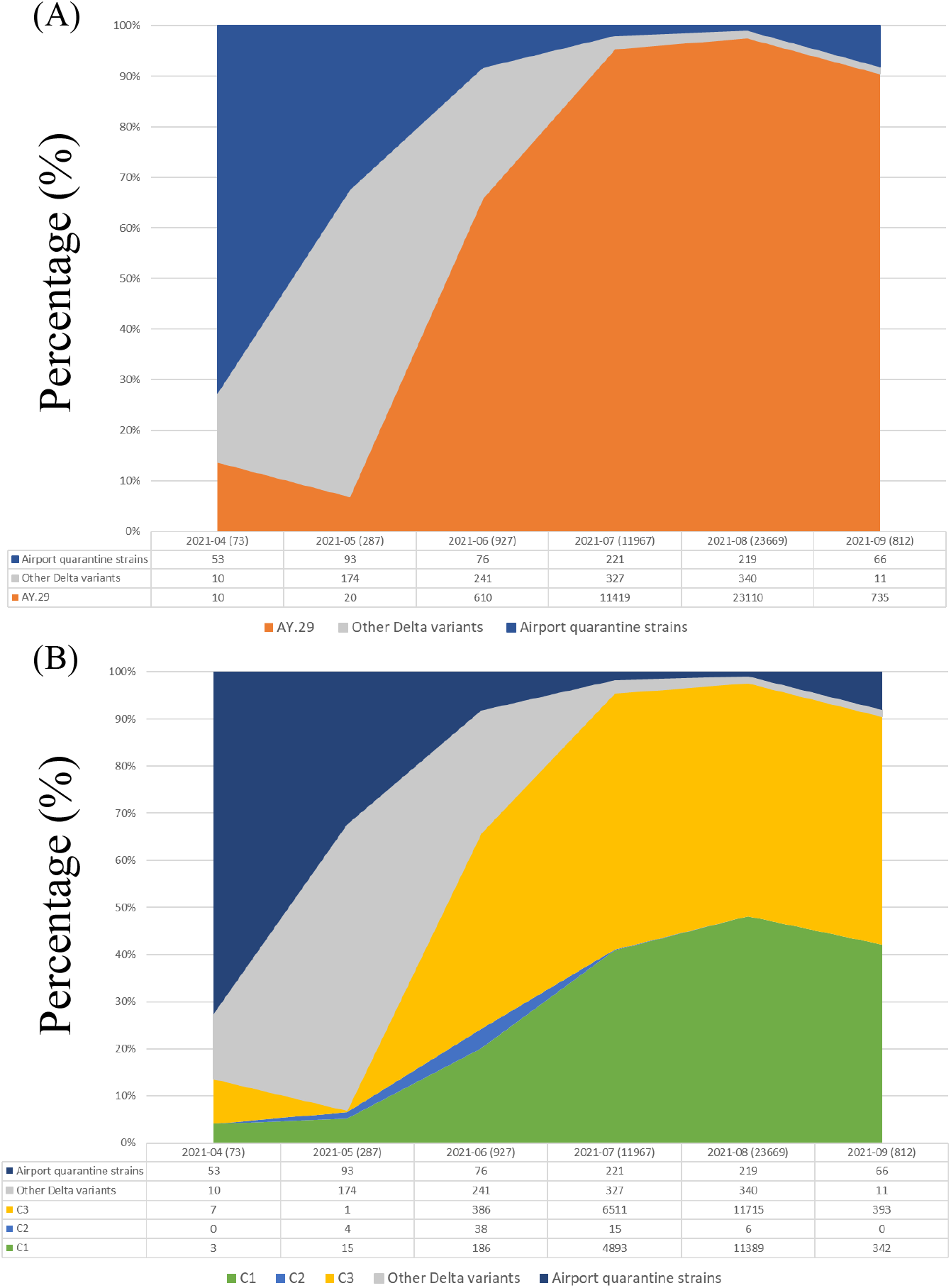
Monthly trends of (A) the AY.29 sub-lineage and (B) subgroups of AY.29 in Japan. The subgruops C1 to C3 are the group IDs defined in Figure 1. The numbers in parentheses are the total numbers of strains isolated in a particular month. The number of strains belonging to each group is shown at the bottom of the figure.

In conclusion, the predominant strain of the Japanese Delta variants currently prevalent in Japan is AY.29, especially between July and September, 2021. In the GISAID data, the number of Delta variants imported from abroad during the Tokyo 2020 Olympics and Paralympics (held in August 2021) was extremely low during the fifth wave in Japan. C5239T and T5514C (ORF1ab: V1750A) are mutation markers that can be used to confirm whether a Delta variant is of Japanese origin. Using these markers, it will be possible to quickly confirm whether new Delta variants originate from AY.29 or from other possibly new variants.

## Data Availability

All data uses the data registered in GISAID and follows the terms of use of GISAID.

## ACKNOWLEDGMENTS

We gratefully acknowledge the authors that have submitted sequences to the GISAID database. We are grateful to Dr. So Nakagawa (Tokai University) for helpful discussions.

